# Trends in diabetes incidence and associated risk factors among people living with HIV in the current treatment era, 2008-2018

**DOI:** 10.1101/2022.04.29.22274506

**Authors:** Gabriel Spieler, Andrew O. Westfall, Dustin M. Long, Andrea Cherrington, Greer A. Burkholder, Nicholas Funderburg, James L. Raper, Edgar T. Overton, Amanda L. Willig

## Abstract

People with HIV (PWH) have an increased risk for diabetes mellitus. Our objectives were to characterize the prevalence and incidence of diabetes in a cohort of people with HIV (PWH), and to evaluate both traditional and HIV-specific risk factors contributing to incident diabetes diagnoses. We conducted a retrospective study of a Southeastern US academic HIV clinic. All PWH age > 18 years of age who attended at least two clinic visits between 2008 and 2018 were evaluated to assess time to diabetes incidence. Laboratory, demographic, clinical, medication and diagnoses data were extracted from the Clinic EMR. Diabetes was defined when at least two of the following three criteria were met: (1) laboratory data consistent with a diagnosis as defined by the ADA SOC (Hgb A1C ≥ 6.5% and/or 2 glucose results >200 mg/dl (at least 30 days apart)), (2) diagnosis of diabetes in the EMR, or (3) exposure to diabetes medication. Time to Diabetes incidence was computed from the entire clinic population for each study year. Univariate Cox proportional hazard models were developed to evaluate associations between each baseline factor and time to DM. Multivariable Cox proportional hazard regression models with time-dependent covariates were created to evaluate the independent association between significant parameters from univariate analyses and time to incident DM. From 4113 PWH included in the analysis, we identified 252 incident cases of diabetes. In multivariable analysis, BMI classification, liver disease, steroid exposure, and use of Integrase Inhibitors were associated with incident diabetes. Additional associated factors included lower CD4 cell counts, duration of HIV infection, exposure to non-statin lipid lowering therapy, and dyslipidemia. Incident diabetes rates are increasing at an alarming rate among PWH. Diabetes prevalence increased from 8.8% in 2008 to 14% in 2018. Both traditional and HIV-related risk factors, particularly body weight, steroid exposure, and use of Integrase Inhibitors, were associated with incident diabetes. Notably, several of the risk factors identified are modifiable and should be targeted for intervention.

## INTRODUCTION

With the significant advances in HIV treatment options, HIV infection has been transformed from a universally fatal disease to a manageable chronic illness.^1^ For those with access to and engagement in HIV care and adherence to ART, certain non-AIDS comorbidities have become more prominent in the management of persons with HIV (PWH).^2^ Insulin resistance and the development of diabetes have become more commonly identified metabolic disturbances.^3, 4^ For many PWH, a combination of structural, socioeconomic, and health issues further contribute to an increased risk of insulin resistance and diabetes mellitus (DM).^2, 3^

Due to the significant morbidity associated with having concomitant HIV infection and diabetes diagnoses, diabetes screening and prevention-focused lifestyle behavior changes recommended for the general population have emerged as important aspects of health management in PWH.^5^ Previous literature supports the hypothesis that both HIV infection and antiretroviral therapy (ART) effects likely contribute to the risk of developing type 2 diabetes. Several mechanisms have been proposed to explain the increased DM risk, including direct effects of viral proteins, persistent systemic inflammation associated with HIV, certain behavioral factors like sedentary lifestyle and poor dietary habits, and medication toxicity.^6-8^ Many early ART agents, notably protease inhibitors, caused dysregulation of glucose homeostasis and increased diabetes risk.^9^ While newer agents are considered metabolically friendly, ART initiation is associated with body composition changes and significant weight gain with worsening of glucose control in at least a subset of patients (REF).^10-12^.

In this analysis, we aimed to characterize the shifting incidence of diabetes in the 1917 HIV Clinic population over time and to evaluate socio-demographic factors, traditional and HIV-specific risk factors for the development of diabetes. We also report prevalence of diabetes by year. By analyzing clinic-wide population trends in the more recent ART era, we can identify key risk factors relevant to today’s PWH and subsequently develop appropriate interventions to reduce the incidence of diabetes and improve the management of diabetes for those who have been diagnosed.

## METHODS

Data were obtained from the University of Alabama at Birmingham (UAB) 1917 HIV/AIDS Clinic Cohort Observational Database Project (UAB 1917 Clinic Cohort). This cohort forms an on-going prospective clinical study, started in 1988, that has collected detailed sociodemographic, psychosocial and clinical data from persons diagnosed with HIV. We conducted a retrospective analysis of 1917 Clinic Cohort data collected between 2008-2018. During this period, the 1917 Clinic used a locally maintained electronic medical record (EMR) which contained detailed provider encounter notes, laboratory values imported from the central UAB laboratory, and electronic prescriptions for all medications. The EMR and study database were quality controlled, with all provider notes reviewed to ensure appropriate data capture regarding diagnoses and medications (including start and stop dates for prescriptions). New and ongoing diagnoses were recorded in patients’ active problem lists. Resolved diagnoses discontinued by the provider remained part of the patient’s EMR after removal from the active problem list. The UAB Institutional Review Board (IRB) approved this study nested in the UAB 1917 Clinic Cohort.

### Participants

Participants with at least two routine care HIV primary provider visits at the UAB 1917 HIV Clinic in Birmingham, AL between January 1, 2008 to December 31, 2018 were included in analysis of HIV incidence. For study inclusion of diabetes incidence, participants additionally were ≥ 18 years of age with a confirmed diagnosis of HIV. Exclusion criteria included no pre-existing DM in the EMR at time of first HIV primary clinic visit, or an extended delay between clinic visits (≥400 days or more after first visit). To report annual prevalence of HIV, the study cohort included all PWH who attended at least one routine care HIV primary provider visits between January 1, 2008 to December 31, 2018.

### Study variables

#### Diabetes diagnosis

Diabetes was defined when at least two of the following three criteria were met: (1) laboratory data consistent with a diagnosis as defined by the ADA SOC (Hgb A1C ≥ 6.5% and/or 2 glucose results >200 mg/dl (at least 30 days apart)),^13^ (2) diagnosis of diabetes in the EMR, or (3) exposure to diabetes medication. Diabetes date of diagnosis was defined for the purposes of this study as the earliest of the following four dates (as documented in the EMR): (1) first diabetes diagnosis, (2) first diabetes medication initiation, (3) first Hgb A1C ≥ 6.5% when a second A1C was also performed, (4) second glucose >200 mg/dl, at least 30 days after first glucose >200 mg/dl. We used nonfasting blood glucose criteria >200 mg/dl versus fasting blood glucose as we could not confirm that all patients in this clinical cohort were fasting at the time of laboratory blood draw.

#### Covariates

The following data elements were abstracted from the EMR: (1) Demographics and vital signs – birth sex, race, ethnicity, birth year, initial clinic visit date, HIV transmission risk factor, insurance status (public, private, uninsured), height (cm), weight (kg), systolic and diastolic blood pressure; (2) Clinical diagnoses as defined within the Center for AIDS Research Network of Integrated Clinical Systems^14^ –hypertension, cardiovascular disease, cerebrovascular disease, renal conditions, malignancies; (3) Prescribed medications: antiretroviral, antihypertensives, statins, testosterone and other hormone replacement therapy, mental health therapies; (4) Laboratory test results: CD4+ T-cell count, HIV viral load, HBV and HCV testing, glucose, hemoglobin A1C, albumin, hemoglobin, triglycerides, LDL-C, HDL-C, total cholesterol, basic chemistries and urine studies. Body mass index (BMI) was computed as weight [kg] / height [m^2^]. The following categories were used to classify patients by BMI: underweight <18.5 kg / m^2^, normal weight 18.5 – 24.9 kg / m^2^, overweight 25 – 29.9 kg / m^2^, obese ≥ 30 kg / m^2^, and Class III obesity ≥ 40 kg / m^2^.

### Statistical Analysis

All data were analyzed using SAS version 9.4 (SAS Institute Inc., Cary, NC, USA).

The primary objective was to determine diabetes incidence and associated risk factors. Three separate analysis were conducted: (1) diabetes incidence by year; (2) time to incident diabetes; and (3) diabetes prevalence by year. Diabetes incidence was computed for the full sample and for each study year as the proportion of patients who developed diabetes in a given year out of the entire clinic population included for analysis for that year. A patient was considered to be in care for a specific calendar year (2008-20018) if they had at least 1 visit during that year. To affirm a DM event, the participant was required to have at least one prior documented visit (i.e. cannot be considered as “incident” at the very first HIV clinic visit). The prior arrived visit could be from an earlier year. Participants with a diabetes diagnosis date before their first arrived visit date in this analysis were excluded. Cochran-Armitage trend test with Agresti-Coull confidence limits was used to evaluate the trend over time.

To determine time to incident diabetes and correlates of diabetes incidence, survivial analyses were conducted beginning with the participant’s first arrived visit and ending with a diabetes diagnosis or censor at the participant’s last arrived HIV clinic visit date. Patients with a gap >400 days between “arrived” visit status at any time were censored at the start of the gap. Multivariable models were fit to assess the association of covariates with diabetes diagnosis. Cox proportional hazards regression models were used to model time to diabetes diagnosis. To account for changes in health status throughout the study period, variables that changed over time were treated as time-varying covariates. Time-varying variables were carried forward up to 365 days (e.g. income, BMI, CD4+, viral load) as some parameters are measured infrequently. Medication exposure variables (ART and non-ART) were considered as time-varying but remain “on” once turned on. Due to previously observed race x sex interactions in diabetes risk, race and birth sex were combined as race/sex categories.

A separate investigation of diabetes prevalence was included to investigate this change over time. For each of the 11 calendar years (2008-2018), a patient was considered “in care” if they had at least one completed HIV clinic visit during that year. Diabetes prevalence for each calendar year was computed as the proportion of participants with a diabetes diagnosis before that year (i.e., diagnosis could occur before 2008) out of the total number of patients in care that year. Change across years was evaluated via Cochran-Armitage trend test with Agresti-Coull confidence limits.

## RESULTS

### Study Population

Of the 5399 PWH who attended one clinic visit or patient orientation with an HIV care provider, 3492 patients met criteria for analysis of diabetes incidence and time to diabetes incidence including diabetes-related laboratory assessment, while 4113 patients met all criteria for analysis of diabetes prevalence. Descriptive characteristics at first clinic visit were not different between the two samples, thus the full sample of 4113 participants are presented in **Table 1**: 871 participants were female (21%), 41 transgender persons (1%); 2554 (62%) Black race, and 1424 (35%) white. At first visit, median age was 38.2 years, median BMI 24.9 kg/m2, median CD4+ count 373 c/mm3, and median HIV viral load was 5700 cp/ml.

**TABLE 1.**
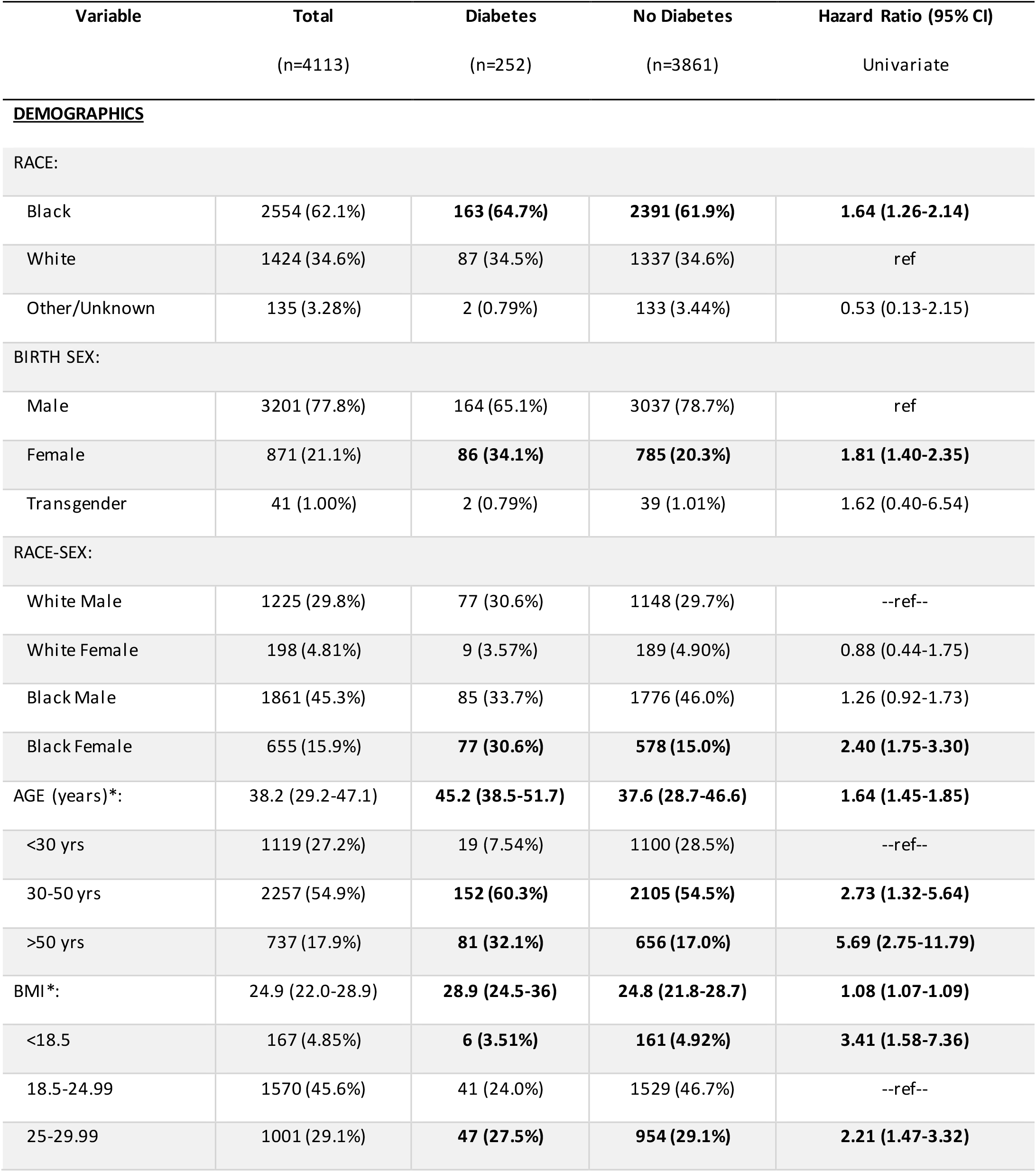
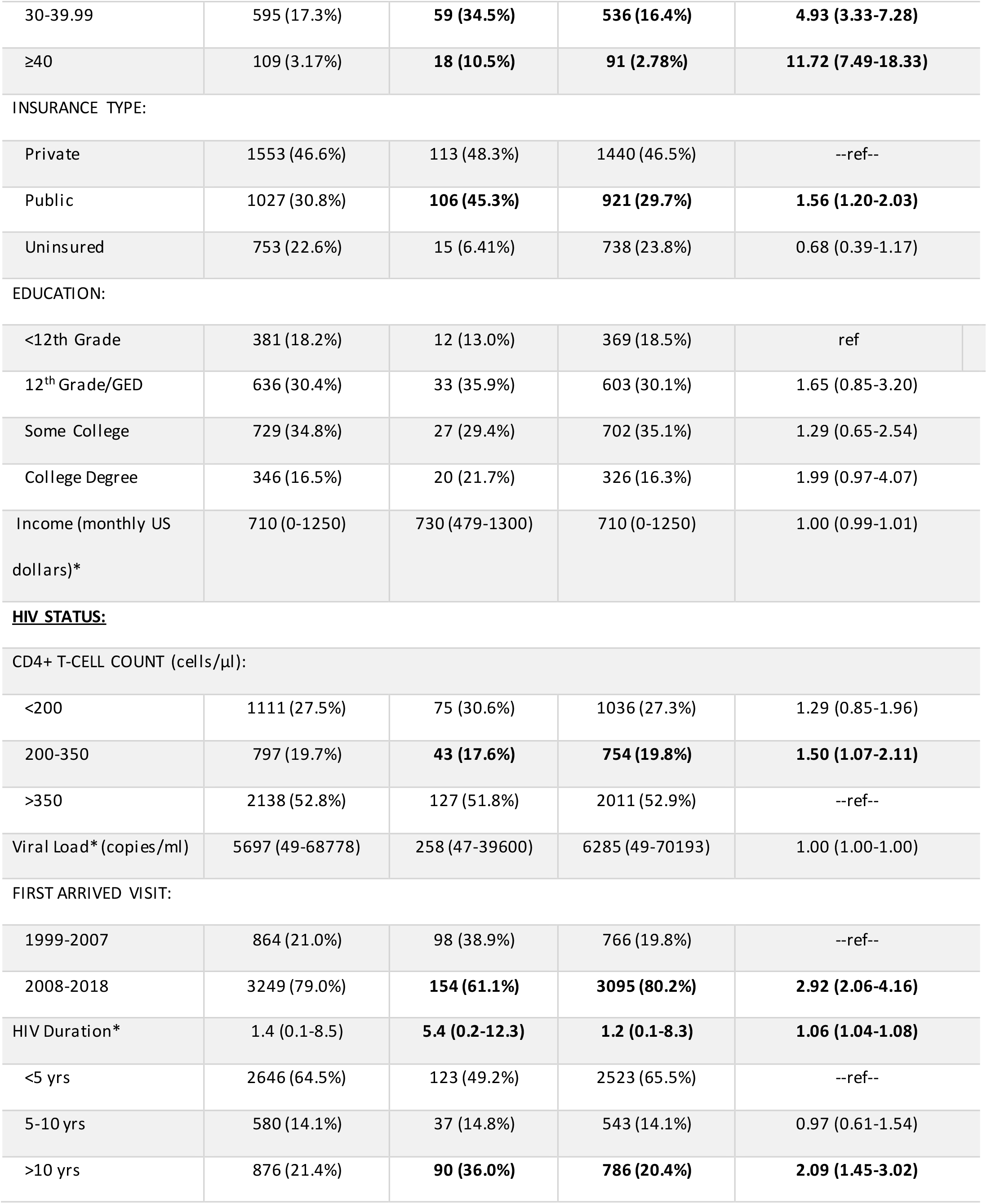
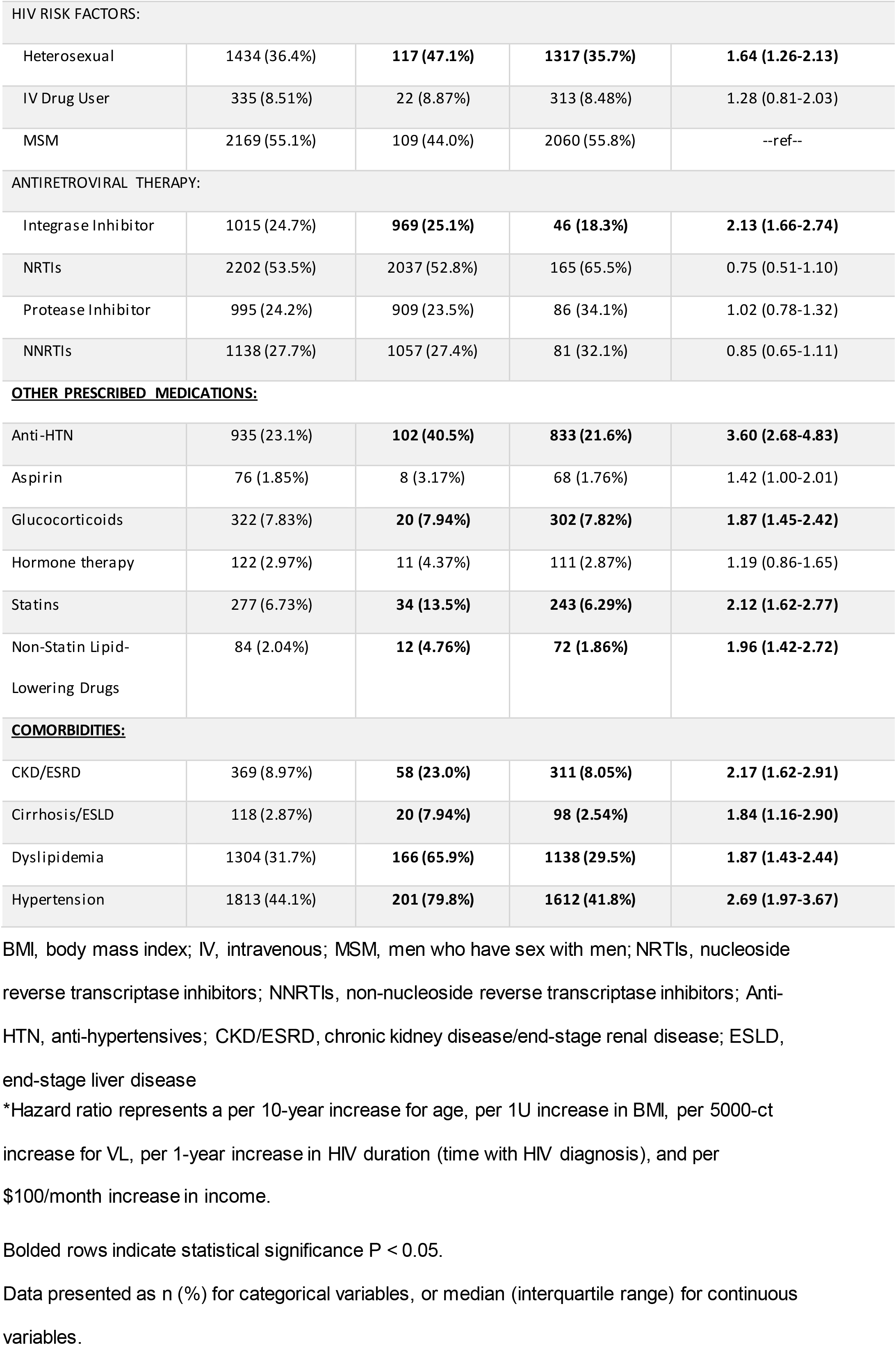
Baseline Characteristics of the Clinic Cohort and Association with Time until Incident Diabetes.

| Variable | Total<br>(n=4113) | Diabetes<br>(n=252) | No Diabetes<br>(n=3861) | Hazard Ratio (95% CI)<br>Univariate |
| --- | --- | --- | --- | --- |
| **DEMOGRAPHICS** |  |  |  |  |
| RACE: |  |  |  |  |
| Black | 2554 (62.1%) | **163 (64.7%)** | **2391 (61.9%)** | **1.64 (1.26-2.14)** |
| White | 1424 (34.6%) | 87 (34.5%) | 1337 (34.6%) | ref |
| Other/Unknown | 135 (3.28%) | 2 (0.79%) | 133 (3.44%) | 0.53 (0.13-2.15) |
| BIRTH SEX: |  |  |  |  |
| Male | 3201 (77.8%) | 164 (65.1%) | 3037 (78.7%) | ref |
| Female | 871 (21.1%) | **86 (34.1%)** | **785 (20.3%)** | **1.81 (1.40-2.35)** |
| Transgender | 41 (1.00%) | 2 (0.79%) | 39 (1.01%) | 1.62 (0.40-6.54) |
| RACE-SEX: |  |  |  |  |
| White Male | 1225 (29.8%) | 77 (30.6%) | 1148 (29.7%) | --ref-- |
| White Female | 198 (4.81%) | 9 (3.57%) | 189 (4.90%) | 0.88 (0.44-1.75) |
| Black Male | 1861 (45.3%) | 85 (33.7%) | 1776 (46.0%) | 1.26 (0.92-1.73) |
| Black Female | 655 (15.9%) | **77 (30.6%)** | **578 (15.0%)** | **2.40 (1.75-3.30)** |
| AGE (years)*: | 38.2 (29.2-47.1) | **45.2 (38.5-51.7)** | **37.6 (28.7-46.6)** | **1.64 (1.45-1.85)** |
| <30 yrs | 1119 (27.2%) | 19 (7.54%) | 1100 (28.5%) | --ref-- |
| 30-50 yrs | 2257 (54.9%) | **152 (60.3%)** | **2105 (54.5%)** | **2.73 (1.32-5.64)** |
| >50 yrs | 737 (17.9%) | **81 (32.1%)** | **656 (17.0%)** | **5.69 (2.75-11.79)** |
| BMI*: | 24.9 (22.0-28.9) | **28.9 (24.5-36)** | **24.8 (21.8-28.7)** | **1.08 (1.07-1.09)** |
| <18.5 | 167 (4.85%) | **6 (3.51%)** | **161 (4.92%)** | **3.41 (1.58-7.36)** |
| 18.5-24.99 | 1570 (45.6%) | 41 (24.0%) | 1529 (46.7%) | --ref-- |
| 25-29.99 | 1001 (29.1%) | **47 (27.5%)** | **954 (29.1%)** | **2.21 (1.47-3.32)** |
| 30-39.99 | 595 (17.3%) | **59 (34.5%)** | **536 (16.4%)** | **4.93 (3.33-7.28)** |
| ≥40 | 109 (3.17%) | **18 (10.5%)** | **91 (2.78%)** | **11.72 (7.49-18.33)** |
| INSURANCE TYPE: |  |  |  |  |
| Private | 1553 (46.6%) | 113 (48.3%) | 1440 (46.5%) | --ref-- |
| Public | 1027 (30.8%) | **106 (45.3%)** | **921 (29.7%)** | **1.56 (1.20-2.03)** |
| Uninsured | 753 (22.6%) | 15 (6.41%) | 738 (23.8%) | 0.68 (0.39-1.17) |
| EDUCATION: |  |  |  |  |
| <12th Grade | 381 (18.2%) | 12 (13.0%) | 369 (18.5%) | ref |
| 12<sup>th</sup> Grade/GED | 636 (30.4%) | 33 (35.9%) | 603 (30.1%) | 1.65 (0.85-3.20) |
| Some College | 729 (34.8%) | 27 (29.4%) | 702 (35.1%) | 1.29 (0.65-2.54) |
| College Degree | 346 (16.5%) | 20 (21.7%) | 326 (16.3%) | 1.99 (0.97-4.07) |
| Income (monthly US dollars)* | 710 (0-1250) | 730 (479-1300) | 710 (0-1250) | 1.00 (0.99-1.01) |
| <200 | 1111 (27.5%) | 75 (30.6%) | 1036 (27.3%) | 1.29 (0.85-1.96) |
| 200-350 | 797 (19.7%) | **43 (17.6%)** | **754 (19.8%)** | **1.50 (1.07-2.11)** |
| >350 | 2138 (52.8%) | 127 (51.8%) | 2011 (52.9%) | --ref-- |
| Viral Load* (copies/ml) | 5697 (49-68778) | 258 (47-39600) | 6285 (49-70193) | 1.00 (1.00-1.00) |
| FIRST ARRIVED VISIT: |  |  |  |  |
| 1999-2007 | 864 (21.0%) | 98 (38.9%) | 766 (19.8%) | --ref-- |
| 2008-2018 | 3249 (79.0%) | **154 (61.1%)** | **3095 (80.2%)** | **2.92 (2.06-4.16)** |
| HIV Duration* | 1.4 (0.1-8.5) | **5.4 (0.2-12.3)** | **1.2 (0.1-8.3)** | **1.06 (1.04-1.08)** |
| <5 yrs | 2646 (64.5%) | 123 (49.2%) | 2523 (65.5%) | --ref-- |
| 5-10 yrs | 580 (14.1%) | 37 (14.8%) | 543 (14.1%) | 0.97 (0.61-1.54) |
| >10 yrs | 876 (21.4%) | **90 (36.0%)** | **786 (20.4%)** | **2.09 (1.45-3.02)** |

| HIV RISK FACTORS: |  |  |  |  |
| --- | --- | --- | --- | --- |
| Heterosexual | 1434 (36.4%) | **117 (47.1%)** | **1317 (35.7%)** | **1.64 (1.26-2.13)** |
| IV Drug User | 335 (8.51%) | 22 (8.87%) | 313 (8.48%) | 1.28 (0.81-2.03) |
| MSM | 2169 (55.1%) | 109 (44.0%) | 2060 (55.8%) | --ref-- |
| ANTIRETROVIRAL THERAPY: |  |  |  |  |
| Integrase Inhibitor | 1015 (24.7%) | **969 (25.1%)** | **46 (18.3%)** | **2.13 (1.66-2.74)** |
| NRTIs | 2202 (53.5%) | 2037 (52.8%) | 165 (65.5%) | 0.75 (0.51-1.10) |
| Protease Inhibitor | 995 (24.2%) | 909 (23.5%) | 86 (34.1%) | 1.02 (0.78-1.32) |
| NNRTIs | 1138 (27.7%) | 1057 (27.4%) | 81 (32.1%) | 0.85 (0.65-1.11) |
| <u>OTHER PRESCRIBED MEDICATIONS:</u> |  |  |  |  |
| Anti-HTN | 935 (23.1%) | **102 (40.5%)** | **833 (21.6%)** | **3.60 (2.68-4.83)** |
| Aspirin | 76 (1.85%) | 8 (3.17%) | 68 (1.76%) | 1.42 (1.00-2.01) |
| Glucocorticoids | 322 (7.83%) | **20 (7.94%)** | **302 (7.82%)** | **1.87 (1.45-2.42)** |
| Hormone therapy | 122 (2.97%) | 11 (4.37%) | 111 (2.87%) | 1.19 (0.86-1.65) |
| Statins | 277 (6.73%) | **34 (13.5%)** | **243 (6.29%)** | **2.12 (1.62-2.77)** |
| Non-Statin Lipid-Lowering Drugs | 84 (2.04%) | **12 (4.76%)** | **72 (1.86%)** | **1.96 (1.42-2.72)** |
| <u>COMORBIDITIES:</u> |  |  |  |  |
| CKD/ESRD | 369 (8.97%) | **58 (23.0%)** | **311 (8.05%)** | **2.17 (1.62-2.91)** |
| Cirrhosis/ESLD | 118 (2.87%) | **20 (7.94%)** | **98 (2.54%)** | **1.84 (1.16-2.90)** |
| Dyslipidemia | 1304 (31.7%) | **166 (65.9%)** | **1138 (29.5%)** | **1.87 (1.43-2.44)** |
| Hypertension | 1813 (44.1%) | **201 (79.8%)** | **1612 (41.8%)** | **2.69 (1.97-3.67)** |

BMI, body mass index; IV, intravenous; MSM, men who have sex with men; NRTIs, nucleoside reverse transcriptase inhibitors; NNRTIs, non-nucleoside reverse transcriptase inhibitors; Anti-HTN, anti-hypertensives; CKD/ESRD, chronic kidney disease/end-stage renal disease; ESLD, end-stage liver disease
\*Hazard ratio represents a per 10-year increase for age, per 1U increase in BMI, per 5000-ct increase for VL, per 1-year increase in HIV duration (time with HIV diagnosis), and per \$100/month increase in income.
Bolded rows indicate statistical significance $P < 0.05$ .
Data presented as n (%) for categorical variables, or median (interquartile range) for continuous variables.

### Diabetes Incidence

Overall, among 3492 participants 252 incident cases were identified in the cohort from a total 28,391 person years (PY) of follow-up. A significant increase in diabetes incidence from 1.04 incidents per 1000 PY, to 1.55 incident per 1000 PY (**Figure 1A**, p< 0.0188). Study participants who developed DM had a median age of 51.9 years old at DM event, compared to an end-of-follow-up-period age of 42.7 years among patients who never developed DM.

**Figure 1a.**
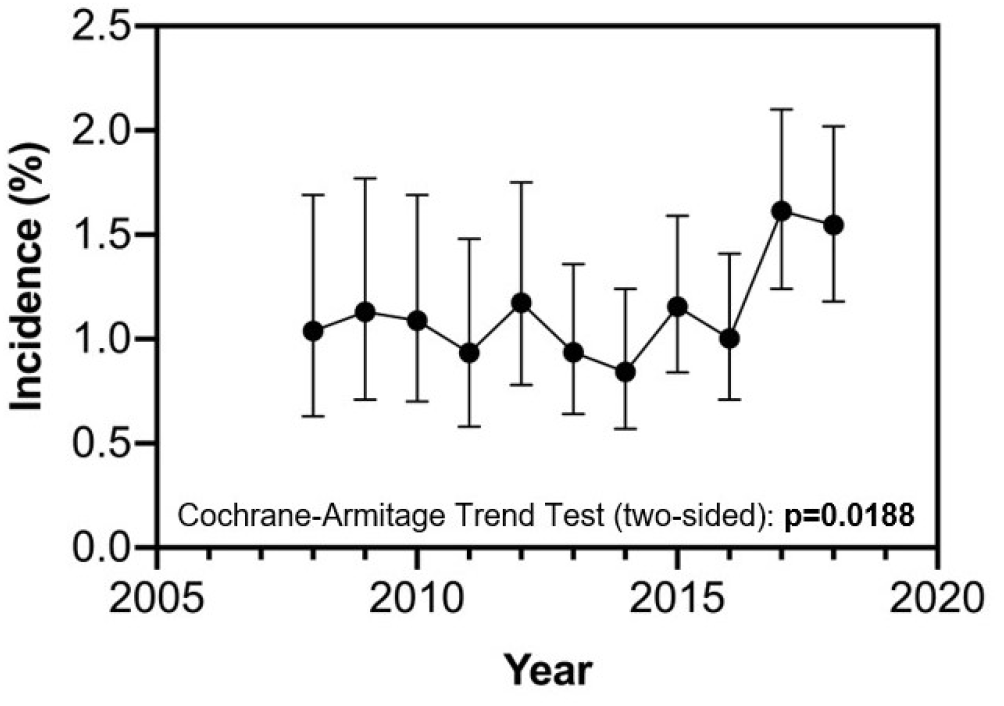
Incident diabetes by year (2008-2018).

**Figure 1b.**
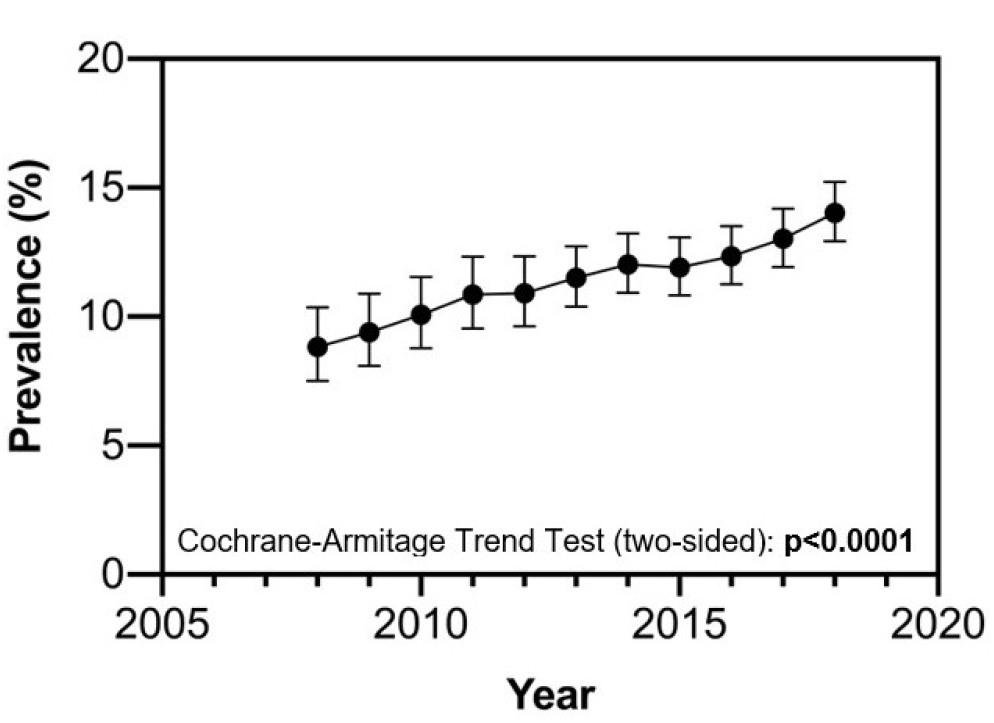
Prevalent diabetes by year (2008-2018).

### Correlates of Diabetes Incidence

In univariate models assessing factors associated with DM diagnosis (**Table 1**), statistically significant associations at *p* < 0.05 were observed with demographic factors including Black race (Hazard Ratio [HR] 1.6), female sex (HR 1.8), Black female race/sex pairing (HR 2.4), older age per 10-year increase (HR 1.6), public insurance (HR 1.6), and first arrived visit between the years of 2008 and 2018 (HR 2.9). Compared to patients with normal weight, incident diabetes was greater when underweight (HR 3.4) and increased across BMI categories, particularly when participants had obesity (HR 4.9) or Class III obesity, i.e. BMI >40kg/m^2^, (HR 11.7). Diabetes diagnosis was also observed with use of several medications at p < 0.05, with the strongest associations for anti-hypertensive exposure (HR 3.6), statins (HR 2.1), non-statin lipid lowering drugs (2.0), and glucocorticoids (1.9). All comorbid conditions evaluated in univariate analyses – kidney disease, liver disease, dyslipidemia, and hypertension - were associated with diabetes incidence. When HIV-specific risk factors were assessed, greater diabetes diagnosis risk was observed in participants with HIV duration post-diagnosis >10 years (HR 2.1), were prescribed integrase inhibitors (HR 2.1), were heterosexual (i.e. did not report MSM, HR 1.6), had CD4+ counts of 200-350 (HR 1.5).

In multivariable analysis (**Figure 2**), risk factors with the strongest associations to diabetes incidence included morbid obesity (adjusted hazard ratio [aHR] 10.5), obesity (aHR 3.9), underweight (aHR 2.8), cirrhosis/end stage liver disease (aHR 1.9), dyslipidemia (aHR 1.6), and HIV duration >10 years (aHR 1.6). Additional risk factors that remained associated with the development of DM in our clinic population were CD4+ counts between 200-350 (aHR 1.55), integrase inhibitor exposure (aHR 1.48), glucocorticoid exposure (aHR 1.46), and non-statin lipid lowering drug exposure (aHR 1.54). Interestingly, Black race/female sex, exposure to statins or anti-hypertensive medications, and hypertension were not associated with diabetes incidence in the modern ART era in multivariable analysis.

**Figure 2.**
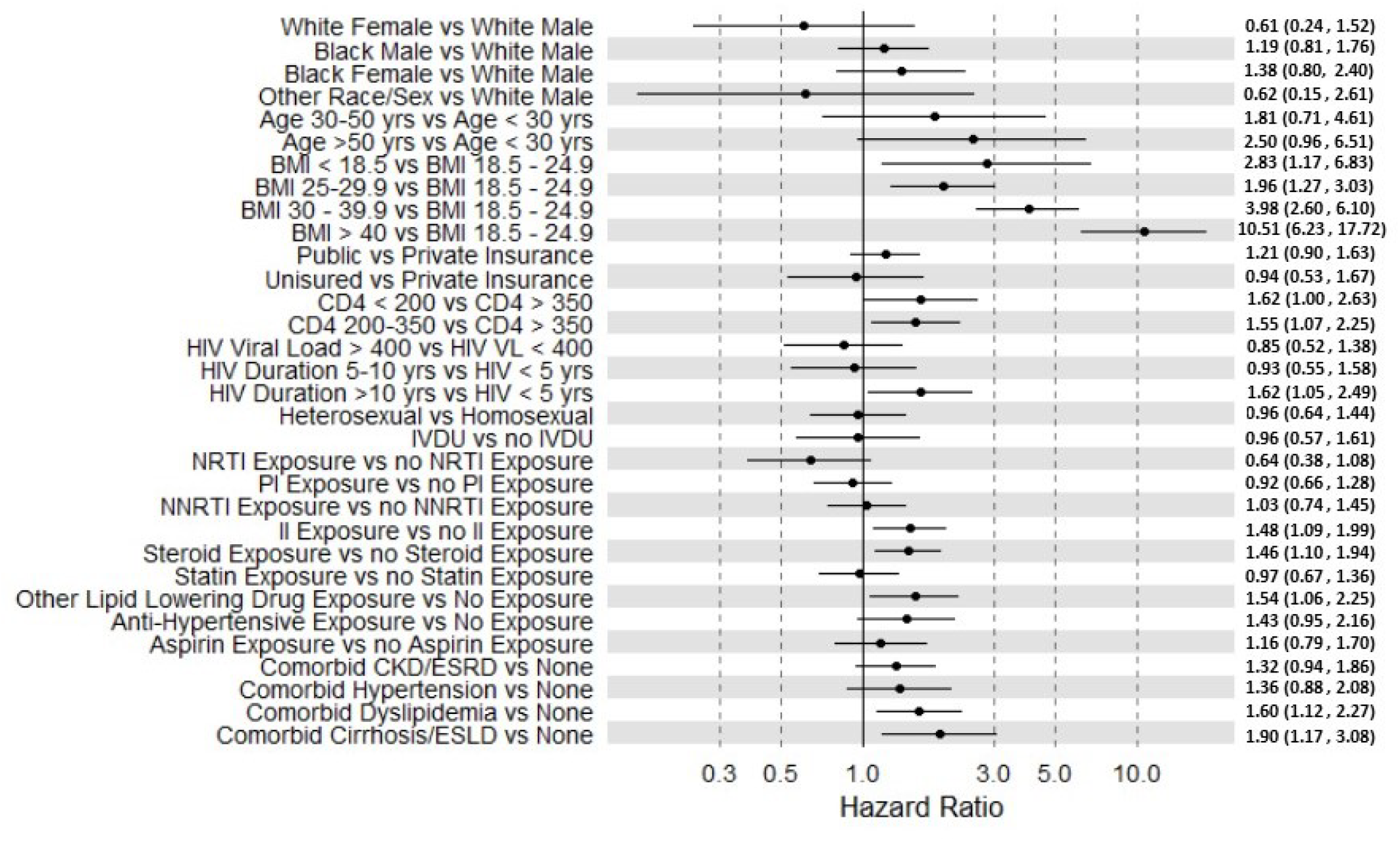
Multivariable analysis of factors associated with increased risk of diabetes diagnosis among people receiving care for HIV between 2008-2018. BMI, body mass index; IV, intravenous; MSM, men who have sex with men; NRTIs, nucleoside reverse transcriptase inhibitors; NNRTIs, non-nucleoside reverse transcriptase inhibitors; II, Integrase Inhibitors; CKD/ESRD, chronic kidney disease/end-stage renal disease; ESLD, end-stage liver disease

### Diabetes Prevalence

We observed an increasing prevalence of diabetes from 8.8% in 2008 to 14.0% in 2018 (Figure 1B, p< 0.001).

## DISCUSSION

Our results show a rapid 10-year increase in incident (to 1.55%) and prevalent (14%) DM among this cohort of PWH from 2008 to 2018. It is unknown whether the rapid increase in diabetes prevalence among this clinic cohort represents a “catch-up” period with the general population in the current treatment era, or whether diabetes prevalence among PWH will surpass HIV un-infected persons. Thus identification of traditional and HIV-related factors that contribute to diabetes risk among PWH is critical to develop prevention and treatment strategies tailored to this high-risk population.

Demographic risk factors have previously been linked to higher risk of developing DM.^3, 15^ We found that persons who self-identify as black females—as well as black race and female sex separately—correlated to a statistically significant increased hazard of developing DM compared to other race/sex groups. However, in the multivariable analysis an independent association was not found to be statistically significant. It is possible that among black females included in our analysis, clustering of other traditional risk factors (e.g. obesity) accounted for this increased risk on the univariate analysis, and a previous study from the 1917 clinic did find that the association of race/sex with a diabetes diagnosis was not observed after adjusting for BMI.^8^ In fact, obesity was the single greatest risk factor associated with the development of DM in our analysis. Moreover, patients with the highest BMIs (>40 kg/m^2^) had the greatest risk of incident DM, with a hazard ratio exceeding 10-fold compared to the referent group (BMI 18.5-24.99 kg/m^2^). At first arrived visit, our data show that 41.4% of patients had a BMI that was at least overweight (BMI >25 kg/m^2^) and 17.1% classified as at least obese (BMI >30 kg/m^2^). This finding confirms previous studies in which higher BMI was associated with diabetes diagnosis and impaired fasting glucose levels.^6, 16, 17^ Given the rising diabetes rates among these PWH and added metabolic risks of HIV infection and ART, more resources should be devoted to curb excess body fat gain and related conditions in this particularly vulnerable population.

Intrinsic biochemical mechanisms of chronic HIV infection as well as ART therapy in DM pathogenesis have been well established,^3, 18^ and are supported by our findings that having HIV for >10 years carries a statistically higher independent risk of developing DM compared to those with HIV for shorter durations. However, the only class of ART independently associated with the development of DM in our analysis were integrase inhibitors, also known as integrase strand transfer inhibitors (INSTIs). Our results are consistent with recent studies, which demonstrate an association between INSTI therapy, diabetes incidence, and weight gain, particularly dolutegravir (DTG).^19, 20^ Given the well-described link between obesity and DM,^21^ it is unsurprising then that INSTI exposure comes with a higher risk of developing DM, at least in part relating to its effects on weight gain. Furthermore, some studies have shown not only an increase in weight gain on INSTIs, but also increased visceral adiposity and truncal obesity, both of which carry increased risks of insulin resistance and DM.^22-24^ This phenomenon may partially account for the link between INSTI exposure and DM risk independent of the other risk factors (e.g. obesity) found in our analysis. Future studies are needed to identify the mechanisms underlying this association, including which INSTI agents are most implicated, and whether the length of time on these agents contributes to which patients subsequently develop diabetes.

Additional risk factors independently associated with the development of DM in our clinic population were older age, lower CD4 counts, longer duration of HIV infection, glucocorticoid exposure, non-statin lipid lowering drug exposure, comorbid cirrhosis/ESLD, and comorbid dyslipidemia. Some factors that were significant in the univariate analysis, including comorbid CKD and statin exposure, were not independently associated with DM risk in the multivariable analysis, and earlier studies have found equivocal results in the association of these variables with diabetes risk in PWH.^25, 26^ This is likely explained by (1) the constellation of multimorbidity and polypharmacy in this study cohort, and (2) limitation of the study period to the most recent treatment decade in which improved HIV treatment options with less viremia-related inflammation and greater obesity prevalence outweigh other factors in diabetes risk.

Results of this study should be interpreted in context of certain limitations. In order to maximize the specificity of the DM group given its importance in this study, fairly stringent criteria were used when defining DM based on patient data from the EMR. Consequently, some patients who actually had DM may have been excluded due to failure to meet those criteria. Additionally, while some patients consistently attended scheduled appointments, others had larger gaps between visits; those with >400 days between visits were excluded from the analysis entirely. If socioeconomic factors impact DM risk, perhaps the least followed patients are also more likely to develop DM but would not be counted as cases of incident diabetes or included in the risk factor analyses. EMR data analysis is limited in the ability to connect prescribed medications, such as statins and glucocorticoids, to specific start/stop dates and diagnoses, particularly when prescribed by healthcare providers outside this EMR hospital system. This limited our ability to evaluate the association of length of exposure for some medications with diabetes risk or to determine the causal factors for association with diabetes incidence in this population. Finally, the 1917 Clinic underwent a significant expansion in patients with increased comorbidities, including diabetes, in 2016-17 following the closure of another local HIV clinic ^27^ which was reflected in the diabetes incidence increase observed subsequently and limited our ability to account for factors including CD4+ nadir. However, no difference in associated risk factors was observed in patients who transferred care during this period compared to the full 1917 Clinic Cohort (data not shown).

PWH have experienced a rapid and clinically meaningful increase in diabetes incidence and prevalence during the current HIV treatment area. Diabetes risk in this population is associated with a combination of HIV-specific (integrase inhibitors) and lifestyle (body fat gain) factors. Diabetes prevention and treatment for PWH must take these factors into account to be effective in reducing the negative health consequences of DM in the setting of HIV.

## Supporting information

Strobe_checklist

## Data Availability

All data produced in the present study are available upon reasonable request to the authors

## Acknowledgements

The authors would like to thank participants of the 1917 Clinic Cohort.

## Authors’ Contributions

Conceived and designed the study: G.S., A.L.W., and E.T.O. Analyzed and interpreted the data: G.S., A.O.W., D.M.L., A.C., G.A.B., N.F., J.L.R., E.T.O, and A.L.W. Drafted the article: G.S., A.O.W., E.T.O., and A.L.W.

## Author Disclosure Statement

No competing financial interests exist.

## Funding Information

This work was supported by the National Institute of Allergy and Infectious Disease at the National Institutes of Health through the UAB Center for AIDS Research [grant number P30-AI27767]. A.C. and A.L.W. were also supported by award numbers P30-DK056336 and P30-DK079626 from the National Institute of Diabetes and Digestive and Kidney Diseases.

## Notes

### Competing Interest Statement

The authors have declared no competing interest.

### Author Declarations

The University of Alabama at Birmingham Institutional Review Board (IRB) approved this study nested in the UAB 1917 Clinic Cohort.

